# Antigenic determinants of SARS-CoV-2-specific CD4^+^ T cell lines reveals M protein-driven dysregulation of interferon signaling

**DOI:** 10.1101/2022.01.20.22269491

**Authors:** Pedro Henrique Gazzinelli-Guimaraes, Gayatri Sanku, Alessandro Sette, Daniela Weiskopf, Paul Schaughency, Justin Lack, Thomas B. Nutman

## Abstract

We generated CD4^+^ T cell lines (TCLs) reactive to either SARSCoV-2 spike (S) or membrane (M) proteins from unexposed naïve T cells from six healthy donor volunteers to understand in fine detail whether the S and M structural proteins have intrinsic differences in driving antigen-specific CD4^+^ T cell responses. Having shown that each of the TCLs were antigen-specific and antigen-reactive, single cell mRNA analyses demonstrated that SARS-CoV-2 S and M proteins drive strikingly distinct molecular signatures. Whereas the S-specific responses are virtually indistinguishable from those responses induced by other viral antigens (e.g. CMV), the M protein-specific CD4^+^ TCLs have a transcriptomic signature that indicate a marked suppression of interferon signaling, characterized by a downregulation of the genes encoding ISG15, IFITM1, IFI6, MX1, STAT1, OAS1, IFI35, IFIT3 and IRF7 (a molecular signature which is not dissimilar to that found in severe COVID-19). Our study suggests a potential link between the antigen specificity of the SARS-CoV-2-reactive CD4^+^ T cells and the development of specific sets of adaptive immune responses. Moreover, the balance between T cells of significantly different specificities may be the key to understand how CD4^+^ T cell dysregulation can determine the clinical outcomes of COVID-19.

## Introduction

Severe acute respiratory syndrome coronavirus 2 (SARS-CoV-2) the novel viral agent of the coronavirus disease 2019 (COVID-19) has resulted in widespread global morbidity and mortality [1]. Infection with SARS-CoV-2 is characterized by a broad spectrum of clinical syndromes, which may range from asymptomatic infection to mild symptoms to severe pneumonia and acute respiratory distress syndrome [2, 3]. According with the World Health Organization weekly epidemiological updates on COVID-19 (Edition 70), the current cumulative number of cases and deaths reported globally is almost 270 million and over 5.3 million respectively.

Immunological and clinical studies of acute and convalescent COVID-19 patients have observed that SARS-CoV-2-specific antibodies and T cell responses are strongly associated with milder disease and accelerated viral clearance [4-6]. Indeed, SARS-CoV-2–specific CD4^+^ and CD8^+^ T cell responses have been reported as crucial for the control and resolution of primary SARS-CoV-2 infection [7]. Varying approaches have been taken to quantify and characterize virus-specific T cell responses in acute, convalescent, and severe patients, in a quest to understand the nature of antigen specificity and function of the adaptive response to SARS-CoV-2 [3, 6, 8, 9]. The response to the structural proteins, including spike (S), nucleocapsid (N), membrane (M), and non-structural proteins (nsp3, nsp4, ORF3a, and ORF8), has been the main targets for study. Using HLA predicted peptide megapools (MP) of the SARS-CoV-2 proteome, Grifoni et al. [10] demonstrated that SARS-CoV-2-specific CD4^+^ T cell responses were found in 100% of patients convalescing from COVID-19, with the majority of the CD4^+^ T cell reactivity directed to SARS-CoV-2 spike, M, and N proteins. On average, these antigens accounted for 27%, 21%, and 11% of the total CD4^+^ T cell response, respectively.

The CD4^+^ T cell responses to the SARS-CoV-2 S or N proteins have been shown to correlate with the magnitude of the anti-SARS-CoV-2 neutralizing antibodies in recovered patients [8, 10], a finding suggesting a potential role for the S protein in triggering a protective response to COVID-19. The SARS-CoV-2 M protein, in contrast, has been implicated in driving evasion of protective immune responses, a process felt to occur by the manipulation of innate antiviral immune responses, most specifically by interfering with interferon (IFN) signaling pathways and by antagonizing the production of type I and III IFN production [11-14]. This M-driven antiviral immune suppression appears to favor SARS CoV-2 viral replication.

To understand in fine detail the molecular nature of the virus-specific CD4^+^ T cell response to the SARS CoV-2 structural proteins, here, we generated (in a non-biased manner) 12 human CD4+ T cell lines (TCLs) reactive to either S protein or M protein from naïve T cells obtained well prior to the COVID19 pandemic from 6 healthy donors with the aim of comparing their molecular properties and function. Our data suggest that SARS-CoV-2 S and M proteins each drive a strikingly distinct molecular signature in TCLs driven under neutral conditions. Whereas the S-specific responses are virtually indistinguishable from those responses induced by other viruses (e.g. CMV), the M protein-specific CD4^+^ TCLs have a transcriptomic signature that demonstrates marked suppression of STAT1-IFRs-interferon pathway signaling, a signature that is virtually indistinguishable from the molecular signature seen associated with severe COVID-19.

## Results

### Specificity and reactivity of CD4^+^ TCLs to SARS-CoV-2 spike and membrane proteins

Purified naïve (CD45RA^+^CD45RO^-^) CD4^+^ T cells from the PBMCs of 6 healthy (unexposed to SARS-CoV-2) donor volunteers, obtained prior to 2019, were differentiated *in vitro* into SARS-CoV-2-specific CD4^+^ TCLs by rounds of stimulation and expansion using autologous dendritic cells loaded with peptide megapools (MP) covering either the entire sequence of SARS-CoV-2 spike protein (MP-S), or covering the complete sequence of SARS-CoV-2 membrane protein (MP-M) (Figure 1A). After three rounds of *in vitro* antigenic stimulation with either S- or M-specific MP in the presence of IL-2 and feeder cells, the CD4^+^ TCLs were assessed. Following *in vitro* stimulation, the generation of the SARS-CoV-2-reactive CD4^+^ T cells to MP-S or MP-M were identified based on the expression of CD154 and CD69. Figure 1B illustrates a representative analysis showing that 44.6% of MP-S-specific CD4^+^ TCL were CD69^+^CD154^+^ in comparison to only 0.87% with their respective naïve CD4^+^ T cells. Similarly, 51.7% of the MP-M-specific CD4^+^ TCL were CD69^+^ and CD154^+^ when compared with 0.59% of their respective naïve CD4^+^ T cells. Moreover, upon stimulation with their respective MP, the vast majority of CD69^+^CD154^+^ MP-S specific- and MP-M specific-CD4^+^ T cell lines were either IFN-y^+^ or TNF-a^+^ (Figure 1B).

**Figure 1:**
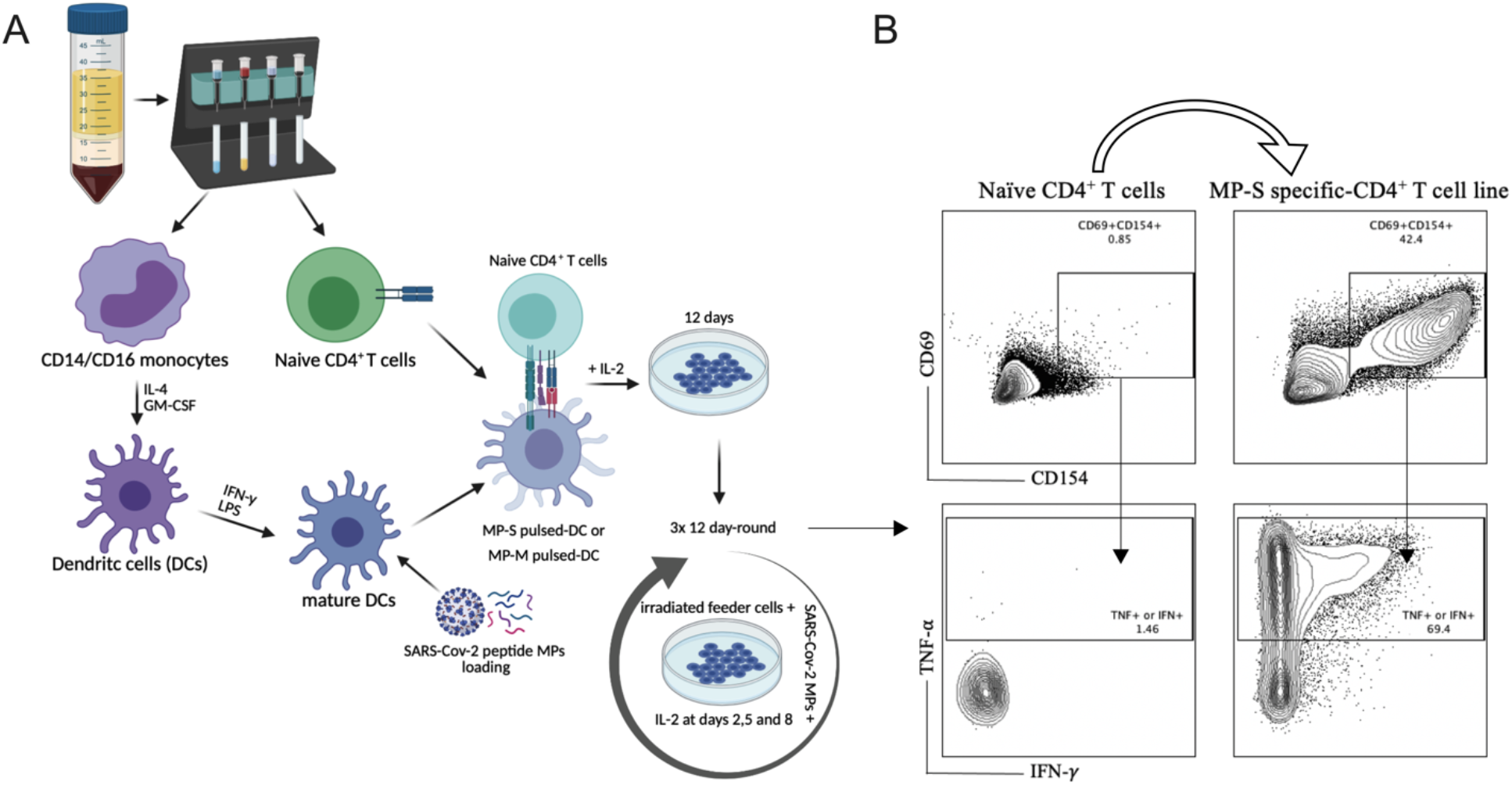
SARS-Cov-2 antigen-specific CD4^+^ T-cell line generation from healthy donor PBMC. Panel A demonstrates the methodology used for generation of the various TCLs beginning with healthy donor naive T-cells and driven by SARS-Cov-2 structural spike (S) and membrane (M) protein-based peptide megapools. Panel B shows a representative flow cytometric analysis of the of MP-S-specific CD4^+^ TCLs expressing CD69^+^CD154^+^ in comparison with their respective naïve CD4^+^ T cells. Moreover, upon stimulation with their respective MPs, the vast majority of CD69^+^CD154^+^ MP-S specific-CD4^+^ T cell lines were either IFN-y^+^ or TNF-a^+^.

We next characterized each of SARS-CoV-2-specific CD4^+^ TCLs for antigen specificity and reactivity (Figure 2). All 6 MP-S specific-(Figure 2A) and MP-M specific-CD4^+^ TCLs (Figure 2B) were cultured *in vitro* in the absence or in the presence of the respective MP or in the presence of a peptide megapool covering the entire sequence of the non-related cytomegalovirus (CMV). PMA/ionomycin was also used as the positive control for the assay. When MP-S-specific CD4^+^ TCLs were stimulated with MP-S, it was observed a significant increase in the frequency in each of the 6 individual CD4^+^ TCLs expressing CD69^+^CD154^+^ and producing either IFN-y^+^ or TNF-a^+^ when compared with non-stimulated cells (24.9 ± 9.5 % vs 5.9 ± 2.3%, p=0.015). This activation failed to occur when the TCLs were stimulated with MP-CMV (8.0 ± 1.9 % vs 5.9 ± 2.3%, p=0.360). Likewise, the stimulation of MP-M-specific CD4^+^ TCLs with MP-M, also induced a marked increase in the frequency in each of the 6 CD4^+^ TCLs expressing CD69^+^CD154^+^ and producing either IFN-y^+^ or TNF-a^+^ when compared with non-stimulated cells (25.4 ± 18.3 % vs 4.3 ± 2.4%, p=0.031). There was little to no reactivity to MP-CMV stimulation (3.3 ± 1.7 % vs 4.3 ± 2.4%, p=0.360) when compared with the media control.

**Figure 2:**
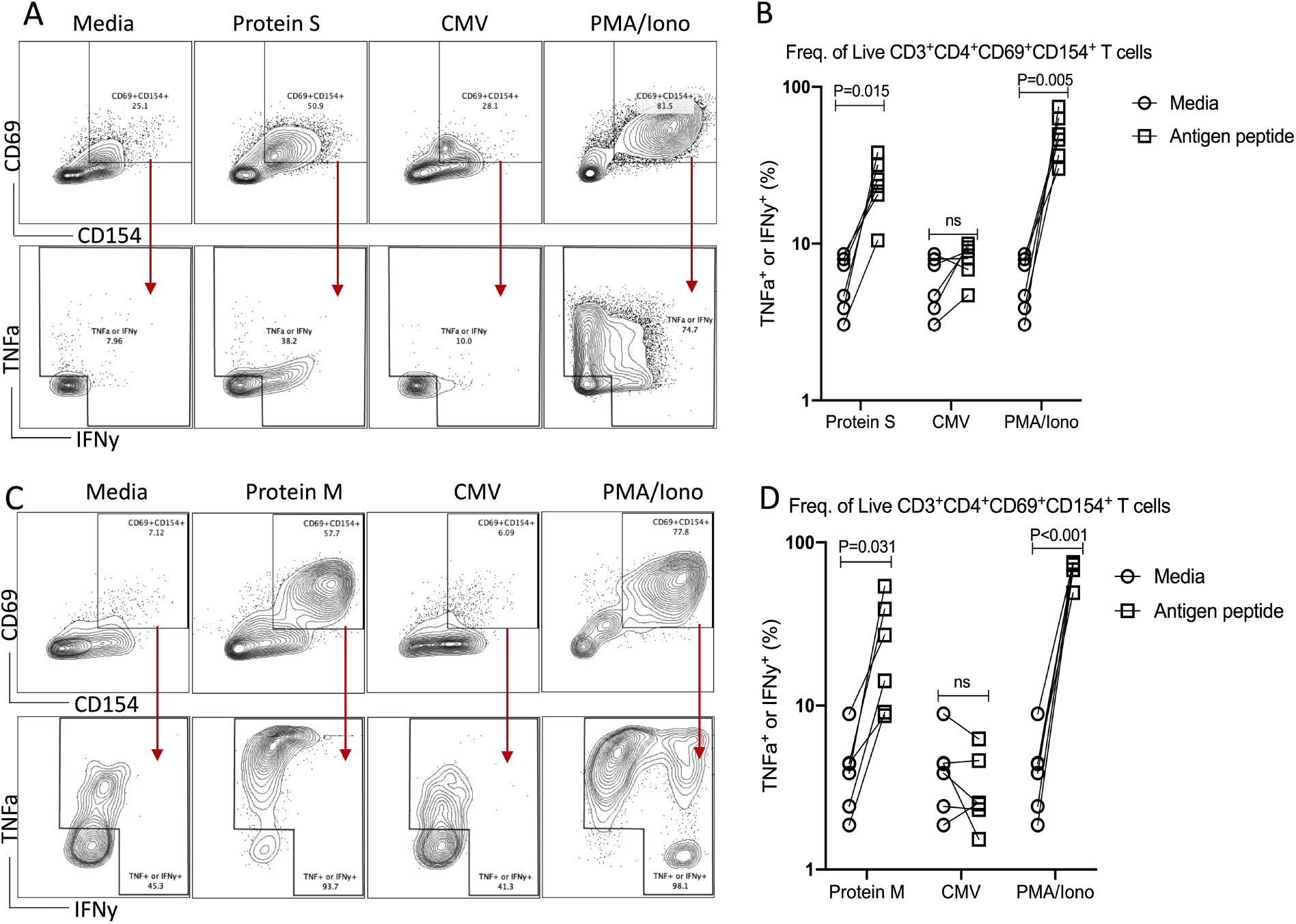
Profiling of SARS-CoV-2 spike (S) and membrane (M) proteins-specific CD4+ T cell line for antigen specificity. SARS-Cov-2 spike protein-specific CD4^+^ TCLs from 6 different donors were cultured *in vitro* in the absence (media) or in the presence of different stimulation conditions including: either SARS-Cov-2 S protein peptide pool (1 µg/peptide) or M protein peptide pool (1 µg/peptide); CMV peptide pool (1µg/peptide); and PMA/ionomycin (0.5/0.05pg/mL). Panels A and C show a representative flow cytometric analysis of the of both MP-S or MP-M-specific CD4^+^ TCLs expressing CD69^+^CD154^+^ and the subsequent antigen-specific cells expressing either IFN-y^+^ or TNF-a^+^ after the different stimulation conditions. Panels B and D reveal the specificity and the reactivity of the six MP-S specific CD4^+^ TCLs (B) or the six MP-M specific CD4^+^ TCLs (D) demonstrating the frequency of CD69^+^CD154^+^ TCLs producing either IFN-y or TNF-a upon stimulation, in comparison with the unstimulated condition. Each plot represents the CD4^+^ TCLs from each donor (n=6) which the circles represent the unstimulated cells and the squares are the respective stimulated conditions. All differences by Wilcoxon matched-pairs test with P < 0.05 are indicated in the graph.

### CD4^+^ TCLs reactive to S and M proteins reveals distinct transcriptomic signature

Having confirmed the specificity and reactivity of each MP-S- and MP-M-specific CD4^+^ TCL, we sorted live CD3^+^CD4^+^ T cells from each TCL (to avoid any contamination with remaining feeder cells or CD8^+^ T cells) and used single cell RNAseq to understand more fully the nature (and heterogeneity) of these TCLs.

Analyses of the single-cell mRNA data of between all the SARS-CoV-2 spike protein-reactive CD4^+^ TCLs with the SARS-Cov-2 membrane protein-reactive CD4+ TCLs revealed a clear distinction and a unique transcriptional signature for each of the M- and S-reactive TCLs (Figure 3A-B). Differential gene expression analysis of the 20 topmost significant genes showed that in contrast to CD4^+^ TCLs reactive to SARS-CoV-2 M protein, the S protein specific-CD4+ T cell lines expressed higher levels of CCL1, GNLY (Granulysin), TNFRSF18 (GITR), NFKBIA, IGLC3, among other genes (Figure 3E), suggesting an effector cytotoxic role for MP-S-reactive CD4^+^ T cells. In marked contrast, CD4^+^ TCLs reactive to SARS-CoV-2 M protein showed increased expression of GZMA (Granzyme A) and the EEF (eukaryotic elongation factors) gene family, including EEF1A1, EEF1B2, EEF1G, EEF2, EIF3E. Most striking, however, the MP-M reactive CD4^+^ T cell lines revealed a marked and significant overall suppressed expression of interferon-inducible genes, including ISG15 (interferon-stimulated gene 15) and IFITM1 (Interferon Induced Transmembrane Protein 1) (Figure 3F).

**Figure 3:**
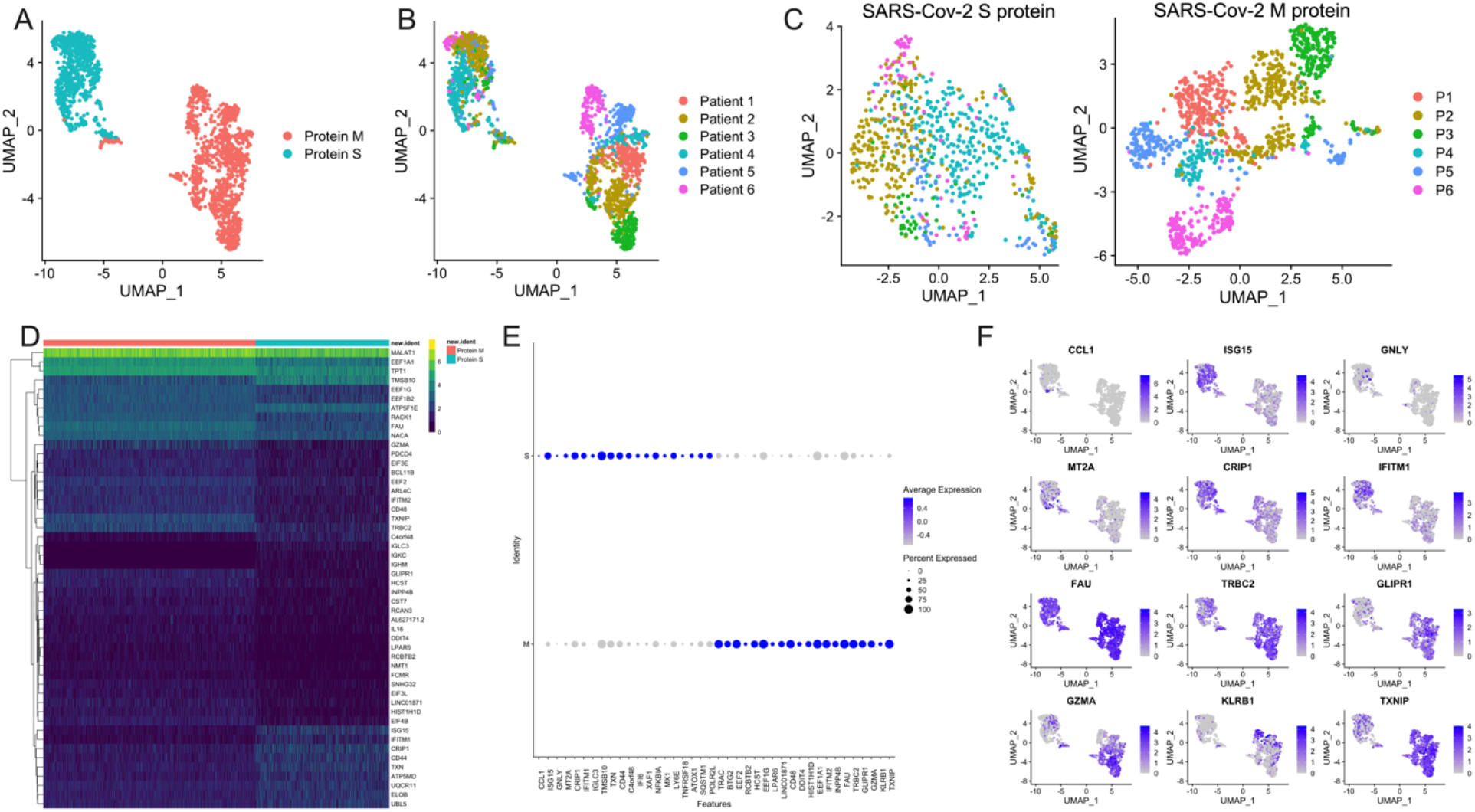
Single-cell transcriptional profiling of SARS-CoV-2 spike (S) and membrane (M) protein-specific CD4^+^ T cell lines (A). Transcriptional signature of both MP-S and MP-M specific-CD4^+^ T cell lines deconvoluted by patient (colored based from P1-P6) is displayed by manifold approximation and projection (UMAP) (B-C). (D) Heatmap showing expression of the most significantly 50 enriched transcripts in each cluster. (E-F) Plot shows average expression and percent expression of selected marker transcripts in each cluster.

Interestingly, comparative analyses between the single-cell transcriptional profiling of SARS-CoV-2 S and M proteins-specific CD4^+^ TCLs and those from CMV-specific-CD4^+^ TCLs (Figure S1A), demonstrated that S-specific responses are virtually indistinguishable from those responses induced by CMV (Figure S1B). Not surprisingly then, the SARS-CoV-2 M-specific responses continue to demonstrate a profoundly marked suppression of the interferon signaling pathway in comparison to CMV-specific CD4^+^ TCLs (Figure S1C), a suppression characterized by a significant downregulation of BAX, IFI6, IFI35, IFIT3, IRF9, ISG15, MX1, OAS1 and STAT1 (Figure S1D).

### SARS-CoV-2 protein M-specific CD4^+^ T cells reveals marked downregulation of interferon pathway signaling

We next performed Ingenuity Pathway Analysis (IPA^®^) on the significantly differentially expressed genes in MP-M-compared with MP-S-reactive CD4^+^ TCLs. Interestingly, the IPA analysis showed that the canonical pathway signaling with immunological relevance most affected in the MP-M-specific CD4^+^ TCLs was the interferon signaling pathway (p=1.59^-10^) and negative z-score (−2.333) (Figure 4A) indicating an extensive and comprehensive suppression of the genes related to types I, II, and III interferon signaling induced by SARS-CoV-2 M protein (Figure 4B), including ISG15, IFITM1, IFI6, MX1, STAT1, OAS1, IFI35, IFIT3 and IRF7 (Figure 4C-D). In addition, upstream analysis also revealed that the MP-M-reactive CD4^+^ TCLs show a marked inhibition of the IFNA2-(interferon alpha 2, z-score= −3.561, p=5.7^-18^), IFNB1-(interferon beta, z-score= −3.312, p=9.6^-14^), IFNG-(interferon gamma, z-score= −2.899, p=7.2^-17^) and IFNL1-(interferon lamba, z-score= −3.646, p=8.3^-17^) associated pathways.

**Figure 4.**
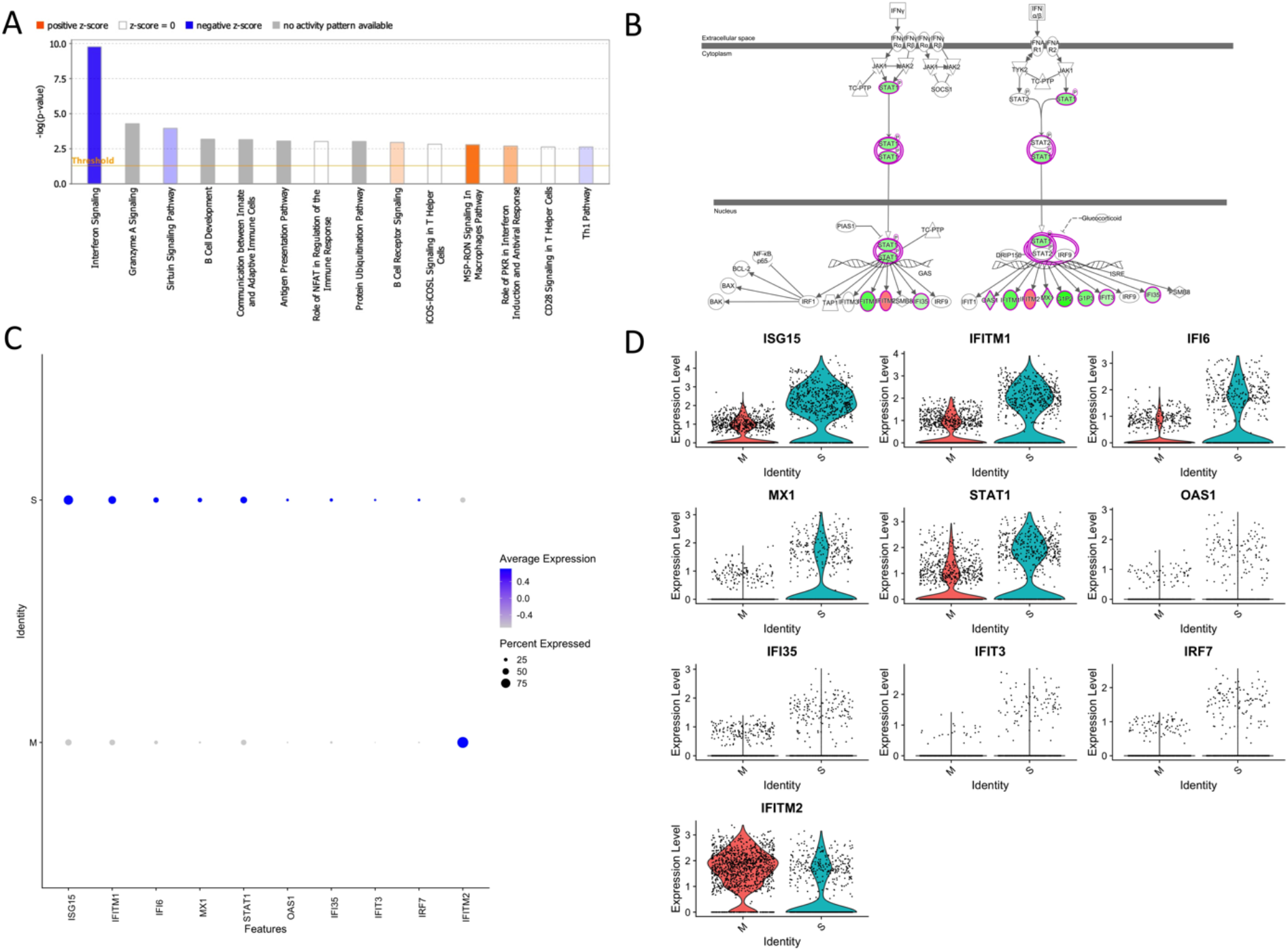
SARS-CoV-2 membrane (M) protein downregulates types I, II, III interferon pathway signaling in CD4+ T cell lines generated from unexposed individuals. Canonical signaling pathways of immunological relevance affected in M protein-specific TCLs (A) indicating a marked suppression interferon signaling pathway and others, including Th1 pathway. (B) Map of the interferon signaling pathway indicating the 9 downstream molecules that were associated with the suppression of interferon, where in green are the genes affected negatively or in red the genes upregulated by SARS-CoV-2 M protein. Feature graph (C) and the violin plot graph (D) showing the average expression and the percent expression of the interferon signaling pathway genes in the respective S-protein-specific CD4^+^ T cell lines and M-protein-specific CD4^+^ T cell lines.

### The molecular signature of SARS-CoV-2 M protein-reactive CD4^+^ TCLs associates with the transcriptional profile seen in severe Covid-19

After characterizing the transcriptional profile of SARS-Cov-2 specific CD4^+^ TCLs, the next step was to use the molecular signature of M protein-reactive T cells to understand how they relate to other diseases and functions through the IPA™ ontology. The transcriptome of SARS-Cov-2 M protein-reactive CD4^+^ TCLs revealed pathways associated with a diverse list of inflammatory diseases and functions, including the top 3: viral infection (107 overlapping molecules, p=7.0^-33^), systemic autoimmune syndrome (86 overlapping molecules, p=1.3^-28^), and most strikingly, Severe COVID-19 (31 overlapping molecules, p=6.8^-28^) (Figure 5B). The 31 severe COVID-19-associated genes that overlapped with the differentially expressed genes in the SARS-Cov-2 M protein-reactive CD4^+^ TCLs were identified and plotted based on their expression level (Figure 5C). Interestingly, the upregulation of the inflammatory genes FOS, JUNB, the downregulation of the interferon-induced genes, including ISG15, IFI6, IFI44, IFIT3, IFI44L, and suppression of the interferon regulatory factors, including (IRF7) driven by M protein were implicated as the key players for this association, suggesting that the molecular signature of SARS-CoV-2 M protein-reactive CD4+ TCLs is characterized by suppression of the same interferon pathways seen in the transcriptional profile of severe Covid-19.

**Figure 5.**
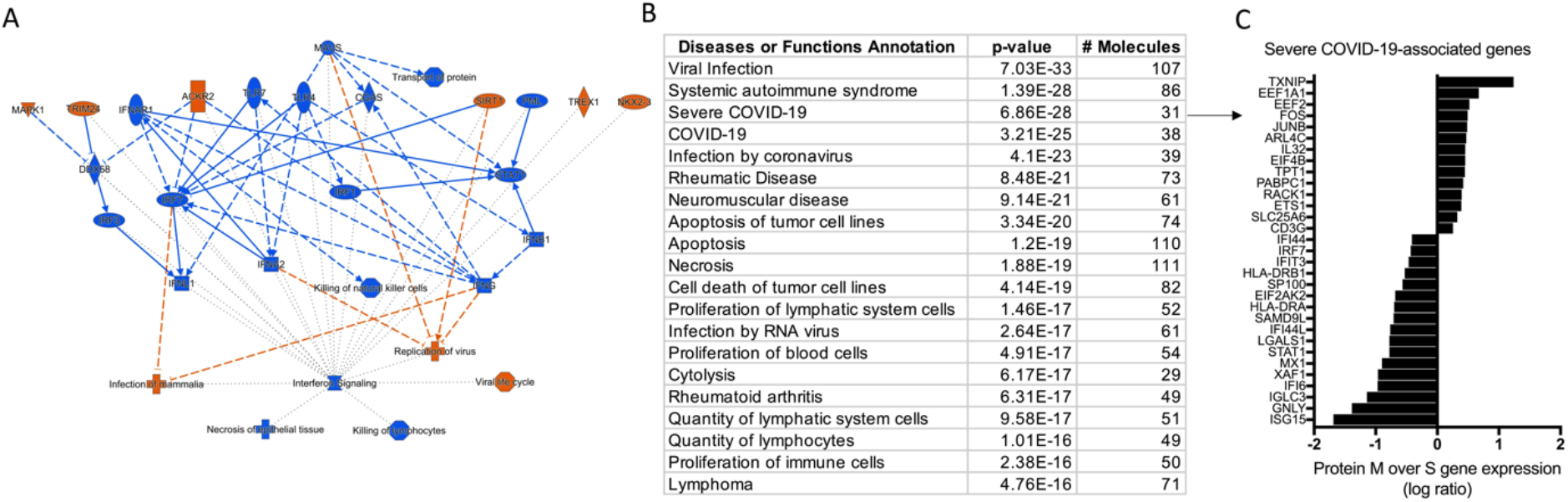
IPA analysis demonstrating an overview summary of the major biological events in the transcriptome of SARS-CoV-2 M protein-reactive CD4^+^ TCLs (A) and The Disease or Function View highlighting how the SARS-Cov-2 specific-CD4^+^ TCLs relate to other diseases and functions through the ingenuity ontology (B). Severe COVID-19 associated genes that overlap with the marked differentially expressed genes in the SARS-Cov-2 M protein-reactive CD4^+^ TCLs (C).

## Discussion

There is a critical need for elucidating the nature of antigen specificity and function of the memory T cell responses to SARS-CoV-2. Understanding the contribution of adaptive immunity to a protective or pathogenic role in SARS-CoV-2 infection may lead the way to a fundamental knowledge that can possibly be used therapeutically in COVID-19 patients or as vaccine targets.

SARS-CoV-2 infected-patients develop specific antibodies, CD4^+^ T cells, and CD8+ T cells in response to the infection [3, 6, 8, 9], although, CD4^+^ T cells had the strongest association with diminished COVID-19 disease severity compared with the other two arms (B cells, CD8+ T cells) of the adaptive immunity [6]. Strikingly, the absence of SARS-CoV-2-specific CD4^+^ T cells was associated with severe or fatal COVID-19 [9]. Data from other another coronavirus (SARS-CoV-1) reported that SARS-CoV-1 spike protein was responsible for nearly two-thirds of the CD4^+^ T cell reactivity with limited reactivity for M and N proteins [15]. It seems, however, that the pattern of antigen predominance of SARS-Cov-2-driven immune responses is different from SARS-CoV-1 in that there is strong reactivity of CD4^+^ T cells to viral S, M and N structural proteins, as well as, to other non-structural proteins and open reading frames, including ORF3 and NSP3 [3, 10, 16-18].

The relationship between antigen-specific CD4^+^ T cell responses and COVID-19 severity remains unclear. First it has been demonstrated that mild COVID-19 patients, who typically recover without special treatment, showed broad SARS-CoV-2-specific CD4^+^ T cell responses to S and N proteins, responses that were highly correlated with specific antibody titers [18]. However, T cell responses were imbalanced in critical ICU patients with a functionally impaired CD4^+^ T cell response showing reduced production of IFN-γ and TNF-α [19].

Indeed, an inflammatory cytokine and chemokine signature (elevated CXCL10, IL-6, and IL-8) accompanied by ineffective interferon responses has been strongly associated with failure to control a primary SARS-CoV-2 infection and with a higher risk of fatal COVID-19 [20-22]. Moreover, impaired, and delayed type I and type III IFN responses have been associated with a higher risk of severe COVID-19 [23]. Interferons (IFNs), including type I (IFN-α and IFN-β) and type III (IFN-λ) are central to both combating virus infection and modulating the antiviral immune response [24, 25]. While type I IFNs are widely expressed and can result in immune-mediated pathology during viral infections, type III IFN (IFN-λ) responses are primarily restricted to mucosal surfaces and are associated with protection to viruses without driving damaging proinflammatory responses [26].

Interestingly, coronaviruses develop efficient immune evasion mechanisms by manipulating immune responses and by interfering with the IFN-related pathways [27]. Indeed, several structural (M and N) and non-structural (NSP1 and NSP3) proteins from SARS-CoV and MERS-CoV can act as interferon antagonists [28]. Notwithstanding, SARS-CoV-2 M protein has also been implicated to antagonize type I and III IFN production by affecting the formation of the RIG-I/MDA-5–MAVS–TRAF3–TBK1 signalosome that has been shown to attenuate antiviral immunity and enhance viral replication [11]. Here, using single-cell transcriptomes of human CD4^+^ TCLs reactive to either S protein or M protein, we have shown that SARS-CoV-2 M protein-reactive CD4^+^ TCLs in comparison with S protein, expressed higher levels of the inflammatory genes FOS, JUNB and lower levels of ISGs, including ISG15, IFI6, IFI35, IFI44, IFIT3, IFITM1, STAT1, OAS1, and interferon regulatory factors, including (IRF7). Viral recognition elicits IFN production, which in turn triggers the transcription of IFN-stimulated genes (ISGs), which engage in various antiviral functions. ISGs have a central role to regulate the type I interferon [29]. Among these ISGs, ubiquitin-like protein ISG15 is one of the most strongly and rapidly induced, and recent work has shown that it can directly inhibit viral replication and modulate host immunity [30-32]. Similarly, we also observed that the molecular signature of SARS-CoV-2 M protein-reactive CD4^+^ TCLs, is characterized by suppression in the interferon pathways, genetically associates with the transcriptional profile of severe Covid-19.

Notably, through signal cell RNAseq analysis of T-cell dysregulation in severe COVID-19 it has been demonstrated that CD4^+^ T cells from severe COVID-19 patients expressed higher levels of a set of inflammatory genes that include FOS, FOSB, JUN and others, gene expression [33] not dissimilar to those found in our M-specific TCLs derived from SARS-CoV-2 unexposed individuals. In parallel, this same study showed that CD4^+^ T cells from patients with severe COVID-19 showed decreased expression of interferon-induced genes including IFIT1, IFIT2, IFIT3, and IFITM1 and those downstream from interferon signaling [33], again striking similar to the molecular signature seen in the M protein-reactive CD4^+^ TCLs in this present study. Finally, the mechanisms by how the peptide megapools of M and S proteins underlies different responses of naïve CD4^+^ T cells remains unclear. Future studies are needed to elucidate if the M-driven dysregulation of interferon signaling pathway in the adaptive immunity resemble to the mechanisms already described including the interaction with pattern recognition receptors (PRRs)-downstream molecules of innate cells, or if it is induced by the interaction of the class II MHC-peptide complex with the restricted TCR repertoire of naïve T cells.

In conclusion, although it has been poorly understood how CD4^+^ T cell dysregulation can contribute to the immunopathogenesis of severe COVID-19, our study suggests a potential link between the antigen specificity of the reactive CD4^+^ T cells to SARS-CoV-2 with the development of a functional and efficient adaptive immune response. The discordant response to S compared to M proteins suggest that the balance between the T cells of different specificities may alter immune evasion mechanisms that may, in turn, drive disease severity. Therefore, one could envision therapeutic approaches that also targets the SARS-CoV-2 M protein may also be important for amelioration of severity of COVID-19.

## Methods

### CMV-specific or SARS-CoV-2 spike or membrane protein-specific CD4^+^ T cell lines generation from unexposed individuals

The protocol for generating an antigen (CMV, SARS-CoV-2-Spike, SARS-CoV-2 Membrane)-specific CD4^+^ T cell line were adapted from previous studies [34, 35]. Briefly, naïve CD4^+^ T cells, as well as monocytes were purified from cryopreserved PBMCs of 6 healthy donor individuals unexposed to SARS-CoV-2 (obtained prior to 2019), using magnetic cell sorting (MACS Miltenyi Biotec, USA). Purified monocytes were differentiated into dendritic cells (DCs) by a 6 day-culture in complete R10 media [RPMI 1640 medium (Gibco) supplemented with 10% heat-inactivated AB serum (Pan Biotech), 1% nonessential amino acids, 1% Hepes 1M, 1 mM sodium pyruvate, 2 mM fresh l-glutamine, 100 μg/ml streptomycin, 100 units/ml penicillin (all from Life Technologies, USA)] at 37°C, 5% CO_2_. 10µL of GM-CSF (1mg/mL) and 10µL of IL-4 (1mg/mL) were added at days 1, 3 and 5. At day 6, immature DCs were harvest, washed and incubated overnight with 10µL of IFN-y (1,000 units/mL) and LPS (100ng/mL) for maturation. At day 7, 1×10^5^ matured DCs were separately seeded in 24 wells plate, and then loaded for 2-4 hours with 1µg/peptide of SARS-CoV-2 peptide megapools (MPs) covering either the entire sequence of SARS-CoV-2 spike protein (MP-S), consisted by a 15-mer peptides overlapping by 10-residues (246 peptides) [10] or covering the complete sequence of SARS-CoV-2 membrane protein (MP-M), consisting of 15-mer sequences with 11 amino acids overlap (PepTivator® SARS-CoV-2 Prot_M, Miltenyi Biotec, USA) and 1µg/peptide of CMV peptide megapools (∼500 peptides). On the top of the either MP-S, MP-M or CMV pulsed-DCs, 1×10^6^ naïve CD4^+^ T cells (1:10 ratio) were added and incubated for 12 days in complete R10 media. On days 2, 5 and 8, 60units of human rIL-2 (Cetus, USA) were added at each well. At day 12, cells in culture were washed, counted, and fed (1:1 ratio) with irradiated (40GY) autologous PBMCs (feeder cells) in the presence of either 1µg/peptide of MP-S or MP-M for another 12-day round culture in the same conditions, including the IL-2 stimulation. After three 12-day rounds of peptide pool stimulation/expansion *in vitro*, we generated a total of 18, 6 MP-S-specific, 6 MP-M-specific and 6 CMV-specific CD4^+^ TCLs, which were profiled for antigen-specificity and reactivity using multiparameter flow cytometry.

### Responsiveness and specificity of SARS-CoV-2-specific CD4^+^ T cell line by flow cytometry

The generation of the different SARS-Cov-2-specific CD4^+^ T cell lines and their specificity and reactivity were confirmed by an immunophenotypic and functional assay where the cell lines were stimulated overnight in 5% CO2 at 37°C with their irradiated autologous feeder cells in the absence (media) or in the presence of their respective antigens (1µg/peptide of MP-S or 1µg/peptide of MP-M). Both CD4^+^ TCLs were also stimulated with 1µg/peptide of a peptide megapool that cover the CD4-predicted epitopes sequences for Cytomegalovirus (MP-CMV) and PMA/ionomycin (Sigma-Aldrich) (0.5/0.05 pg/ml). The cells were then stained for viability (Live/Dead fixable blue (UV450), Molecular Probes), and then incubated with anti-CD3 (BV421), anti-CD4 (PerCP-Cy5.5) for 30 min in the dark at room temperature. The cells were next washed twice with FACS buffer, then fixed and permeabilized using a Fix/Perm buffer kit (BioLegend) for 30 min at 4°C. The cells were washed twice with Perm buffer (BioLegend) and resuspended with the intracellular antibody pool containing anti-CD69 (FITC), anti-CD154 (APC), anti-TNF-a (Alexa Fluor 700) and anti-IFN-y (BUV737) (Supplemental Table I) for 30 min at 4°C. Finally, the cells were washed twice with Perm buffer and then acquired using the BD LSRFortessa flow cytometer (BD Biosciences) and FACSDiva software (BD Biosciences) for acquisition. All analyses were performed using FlowJo v10.5.3.

### DNA purification for whole exome sequencing

Genomic DNA purified (Promega) from 30×10^6^ PBMCs of the 6 healthy donor individuals unexposed to SARS-CoV-2 (the donors we used to generate the SARS-CoV-2-specific CD4+ TCLs) was sent for whole exome sequencing and HLA-typing at Psomagen, Inc.

### Sample preparation and single-cell RNA-seq libraries for next generation sequencing

In summary, 1×10^5^ live CD3^+^CD4^+^ T cells were sorted from the 6 MP-S, MP-M and CMV-specific CD4^+^ T cell lines cultures using a BD FACSAria Cell Sorter (BD Biosciences). 1 × 10^4^ cells of each CD4^+^ TCLs were pooled together for both conditions separately. Approximately 6 × 10^3^ multiplexed cells (a thousand cells per donor for each cell line) were loaded in three lanes of the Chromium Next GEM Chip G (10x Genomics) respectively, one for MP-S, another for MP-M and another for CMV-specific-CD4^+^ TCLs, resulting in three 10x Genomics Single Cell Chromium 3’ mRNA libraries, made in accordance with Chromium Single Cell 3’ Reagent Kits User Guide (v3.1).

### Single cell RNA-Seq analysis

Two 10x Genomics Single Cell Chromium 3’ mRNA libraries were made and sequenced as part of one Illumina NextSeq run. Each sample had a sequencing yield of greater than 68 million reads. The sequencing run was setup with 150 cycles + 150 cycles symmetric run. Initial processing of the two samples included removal of the cells with extremely low number of UMI counts using Cellranger v4.0.0 using default parameters except for the forced cell counts which were 1,979 for S and 1,461 for M. Demuxlet [36] was used with matching exome SNP data to call cell genotypes and multiplet annotations. The remainder of the single cell RNA-Seq (scRNA) analysis was performed with Seurat v3.2.2 [37]. SingleR [38] utilizing data available from the Novershtern Hematopoietic database [39] was used for cell type identification. For sample S, after filtering for less than 500 and greater than 6000 detected genes, higher than 15% mitochondrial gene expression, anything other than CD4+ annotation by SingleR, and multiplets called by demuxlet, retained 713 of the 1,979 cells. For Sample M, after filtering for less than 200 and greater than 5000 detected genes, higher than 15% mitochondrial gene expression, anything other than CD4^+^ annotation by SingleR, and multiplets called by demuxlet, retained 1,132 of the 1,461 cells. Samples S and M were merged using Seurat and standard scRNA analysis was done using 20 principal components (PCs) to visualize the UMAP diagrams. Initial clustering with UMAP identified 2 cell clusters separating the S and M samples. Differential expression analysis was performed to identify cluster-specific markers and to compare the S and M cell populations using MAST [40], or “Model-based Analysis of Single-cell Transcriptomics”, whereas the cluster-specific canonical pathway enrichment profiles were generated using Ingenuity Pathway Analysis (IPA, Qiagen, Redwood City, CA, USA).

### Study approval

This study (NCT00001230) was approved by the National Institute of Allergy and Infectious Diseases (NIAID) Institutional Review Board. Written informed consent was obtained from all participants.

## Data Availability

All data produced in the present work are contained in the manuscript

## Author contributions

Conceptualization: PHGG, GS, AS, DW and TBN. Methodology: PHGG, GS, AS, DW, PS, JL. Investigation: PHGG, GS, PS, JL. Visualization: PHGG, GS, PS. Funding acquisition: PHGG, TBN. Project administration: PHGG, TBN. Supervision: PHGG, TBN. Writing – original draft: PHGG. Writing – review & editing: PHGG, GS, AS, DW, PS, JL, TBN.

## Acknowledgments

This study was supported by the Division of Intramural Research, NIAID, NIH and partly by NIH contract Nr. 75N9301900065 (D.W., A.S.).

## References

1. Hu, B., et al., Characteristics of SARS-CoV-2 and COVID-19. Nat Rev Microbiol, 2021. 19(3): p. 141–154.

2. Raoult, D., et al., Coronavirus infections: Epidemiological, clinical and immunological features and hypotheses. Cell Stress, 2020. 4(4): p. 66–75.

3. Le Bert, N., et al., SARS-CoV-2-specific T cell immunity in cases of COVID-19 and SARS, and uninfected controls. Nature, 2020. 584(7821): p. 457–462.

4. Tan, A.T., et al., Early induction of functional SARS-CoV-2-specific T cells associates with rapid viral clearance and mild disease in COVID-19 patients. Cell Rep, 2021. 34(6): p. 108728.

5. Liao, M., et al., Single-cell landscape of bronchoalveolar immune cells in patients with COVID-19. Nat Med, 2020. 26(6): p. 842–844.

6. Rydyznski Moderbacher, C., et al., Antigen-Specific Adaptive Immunity to SARS-CoV-2 in Acute COVID-19 and Associations with Age and Disease Severity. Cell, 2020. 183(4): p. 996–1012 e19.

7. Dan, J.M., et al., Immunological memory to SARS-CoV-2 assessed for up to eight months after infection. bioRxiv, 2020.

8. Ni, L., et al., Detection of SARS-CoV-2-Specific Humoral and Cellular Immunity in COVID-19 Convalescent Individuals. Immunity, 2020. 52(6): p. 971–977 e3.

9. Sette, A. and S. Crotty, Adaptive immunity to SARS-CoV-2 and COVID-19. Cell, 2021. 184(4): p. 861–880.

10. Grifoni, A., et al., Targets of T Cell Responses to SARS-CoV-2 Coronavirus in Humans with COVID-19 Disease and Unexposed Individuals. Cell, 2020. 181(7): p. 1489–1501 e15.

11. Zheng, Y., et al., Severe acute respiratory syndrome coronavirus 2 (SARS-CoV-2) membrane (M) protein inhibits type I and III interferon production by targeting RIG-I/MDA-5 signaling. Signal Transduct Target Ther, 2020. 5(1): p. 299.

12. Siu, K.L., et al., Severe acute respiratory syndrome coronavirus M protein inhibits type I interferon production by impeding the formation of TRAF3.TANK.TBK1/IKKepsilon complex. J Biol Chem, 2009. 284(24): p. 16202–16209.

13. Xia, H., et al., Evasion of Type I Interferon by SARS-CoV-2. Cell Rep, 2020. 33(1): p. 108234.

14. Fu, Y.Z., et al., SARS-CoV-2 membrane glycoprotein M antagonizes the MAVS-mediated innate antiviral response. Cell Mol Immunol, 2021. 18(3): p. 613–620.

15. Li, C.K., et al., T cell responses to whole SARS coronavirus in humans. J Immunol, 2008. 181(8): p. 5490–500.

16. Nelde, A., et al., SARS-CoV-2-derived peptides define heterologous and COVID-19-induced T cell recognition. Nat Immunol, 2021. 22(1): p. 74–85.

17. Peng, Y., et al., Broad and strong memory CD4(+) and CD8(+) T cells induced by SARS-CoV-2 in UK convalescent individuals following COVID-19. Nat Immunol, 2020. 21(11): p. 1336–1345.

18. Oja, A.E., et al., Divergent SARS-CoV-2-specific T- and B-cell responses in severe but not mild COVID-19 patients. Eur J Immunol, 2020. 50(12): p. 1998–2012.

19. Zheng, H.Y., et al., Elevated exhaustion levels and reduced functional diversity of T cells in peripheral blood may predict severe progression in COVID-19 patients. Cell Mol Immunol, 2020. 17(5): p. 541–543.

20. Aid, M., et al., Vascular Disease and Thrombosis in SARS-CoV-2-Infected Rhesus Macaques. Cell, 2020. 183(5): p. 1354–1366 e13.

21. Kuri-Cervantes, L., et al., Comprehensive mapping of immune perturbations associated with severe COVID-19. Sci Immunol, 2020. 5(49).

22. Del Valle, D.M., et al., An inflammatory cytokine signature predicts COVID-19 severity and survival. Nat Med, 2020. 26(10): p. 1636–1643.

23. Kim, Y.M. and E.C. Shin, Type I and III interferon responses in SARS-CoV-2 infection. Exp Mol Med, 2021. 53(5): p. 750–760.

24. Muller, U., et al., Functional role of type I and type II interferons in antiviral defense. Science, 1994. 264(5167): p. 1918–21.

25. Lee, A.J. and A.A. Ashkar, The Dual Nature of Type I and Type II Interferons. Front Immunol, 2018. 9: p. 2061.

26. Hadjadj, J., et al., Impaired type I interferon activity and inflammatory responses in severe COVID-19 patients. Science, 2020. 369(6504): p. 718–724.

27. Taefehshokr, N., et al., Covid-19: Perspectives on Innate Immune Evasion. Front Immunol, 2020. 11: p. 580641.

28. Fung, T.S. and D.X. Liu, Human Coronavirus: Host-Pathogen Interaction. Annu Rev Microbiol, 2019. 73: p. 529–557.

29. Schneider, W.M., M.D. Chevillotte, and C.M. Rice, Interferon-stimulated genes: a complex web of host defenses. Annu Rev Immunol, 2014. 32: p. 513–45.

30. Der, S.D., et al., Identification of genes differentially regulated by interferon alpha, beta, or gamma using oligonucleotide arrays. Proc Natl Acad Sci U S A, 1998. 95(26): p. 15623–8.

31. Loeb, K.R. and A.L. Haas, The interferon-inducible 15-kDa ubiquitin homolog conjugates to intracellular proteins. J Biol Chem, 1992. 267(11): p. 7806–13.

32. Perng, Y.C. and D.J. Lenschow, ISG15 in antiviral immunity and beyond. Nat Rev Microbiol, 2018. 16(7): p. 423–439.

33. Kalfaoglu, B., et al., T-Cell Hyperactivation and Paralysis in Severe COVID-19 Infection Revealed by Single-Cell Analysis. Front Immunol, 2020. 11: p. 589380.

34. Nutman, T.B., et al., Parasite antigen-specific human T cell lines and clones. Major histocompatibility complex restriction and B cell helper function. J Clin Invest, 1984. 73(6): p. 1754–62.

35. Kahlert, H., Production of T-cell lines. Methods Mol Med, 2008. 138: p. 31–41.

36. Kang, H.M., et al., Multiplexed droplet single-cell RNA-sequencing using natural genetic variation. Nat Biotechnol, 2018. 36(1): p. 89–94.

37. Stuart, T., et al., Comprehensive Integration of Single-Cell Data. Cell, 2019. 177(7): p. 1888–1902 e21.

38. Aran, D., et al., Reference-based analysis of lung single-cell sequencing reveals a transitional profibrotic macrophage. Nat Immunol, 2019. 20(2): p. 163–172.

39. Novershtern, N., et al., Densely interconnected transcriptional circuits control cell states in human hematopoiesis. Cell, 2011. 144(2): p. 296–309.

40. Finak, G., et al., MAST: a flexible statistical framework for assessing transcriptional changes and characterizing heterogeneity in single-cell RNA sequencing data. Genome Biol, 2015. 16: p. 278.

