## Supplemental Figure 1 for "Antigenic determinants of SARS-CoV-2-specific CD4^+^ T cell lines reveals M protein-driven dysregulation of interferon signaling"

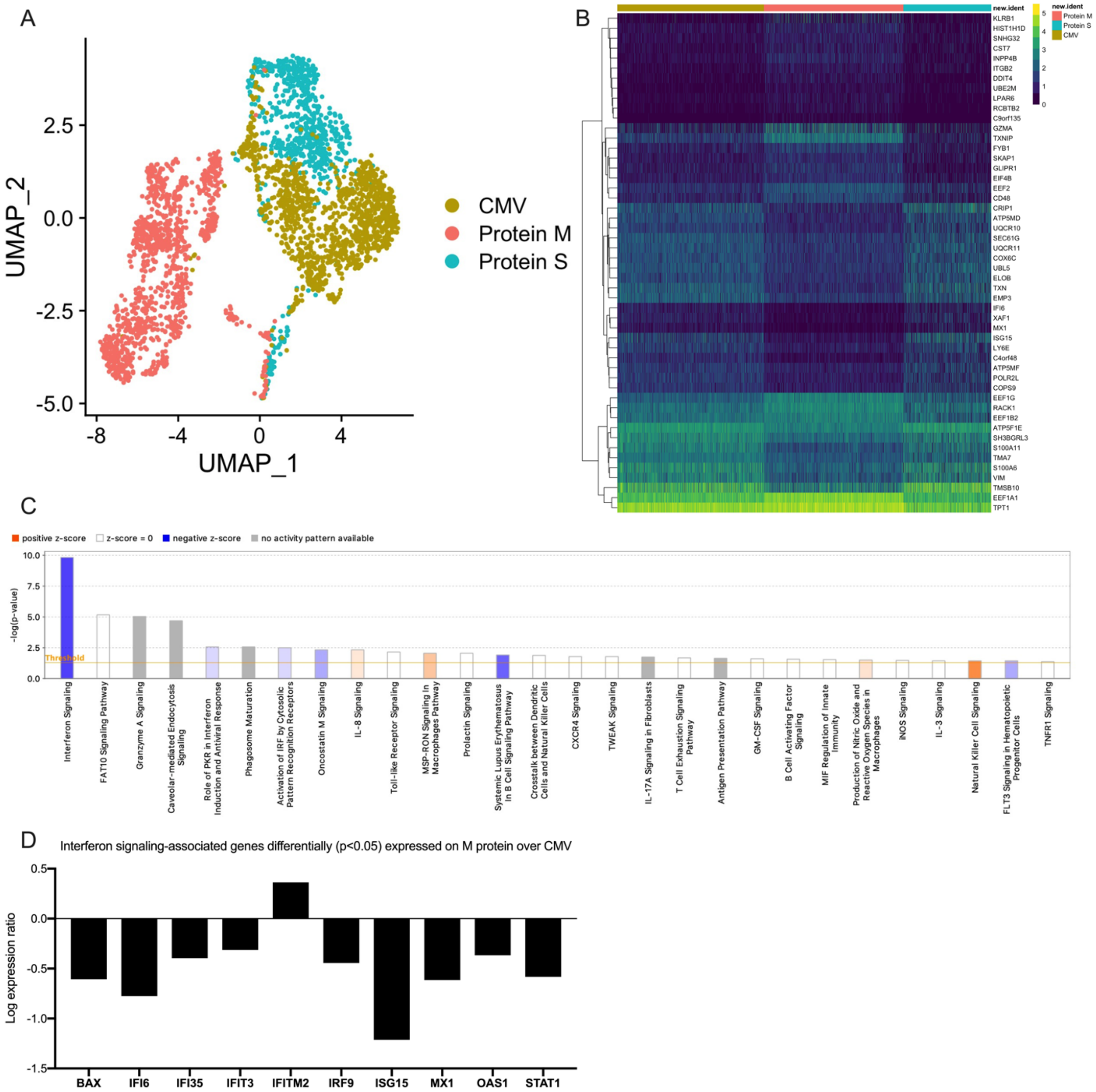


**Supplemental figure 1.** Single-cell transcriptional profiling of SARS-CoV-2 S, M and CMV-specific CD4$^+$ T cells (A). Heatmap showing expression of the most significantly 50 enriched transcripts in M protein-specific CD4$^+$ T cell lines over S protein- and CMV-specific CD4$^+$ T cell lines (B). Canonical signaling pathways (IPA, QIAGEN) of immunological relevance affected in M protein-specific TCLs over CMV-specific TLCs indicating a marked suppression of interferon signaling pathway (C), with the associated gene expression levels and directions presented individually (D).
